# RFA-U-Net: A Foundation Model-Driven Approach for Accurate Choroid Segmentation in OCT Imaging

**DOI:** 10.1101/2025.05.03.25326923

**Authors:** Alireza Hayati, Roya Arian, Narges Saeedizadeh, William Rojas, Rupesh Agrawal, Raheleh Kafieh

**Affiliations:** Students’ Research Committee (SRC), Qazvin University of Medical Sciences, Qazvin, Iran; Department of Engineering, Durham University, South Road, Durham, UK; Institute for Intelligent Systems Research and Innovation, Deakin University, Geelong, Australia; Programme for Ocular Inflammation and Infection Translational Research, National Healthcare Group Eye Institute, Singapore, Singapore; Lee Kong Chian School of Medicine, Nanyang Technological University of Singapore, Singapore; Tan Tock Seng Hospital, National Healthcare Group Eye Institute, Singapore, Singapore; Singapore Eye Research Institute, Singapore; Duke NUS Medical School, Singapore, Singapore

**Keywords:** Segmentation, Transformer, Choroid Segmentation, OCT Images, Foundation Model

## Abstract

The choroid layer plays a critical role in maintaining outer retinal health and is implicated in numerous vision-threatening diseases such as diabetic retinopathy, age-related macular degeneration, and diabetic macular edema. While Enhanced-Depth Imaging Optical Coherence Tomography (EDI-OCT) enables detailed visualization of the choroid, accurate manual segmentation remains time-consuming and subjective. In this study, we introduce RFA-U-Net, a novel deep learning model designed for precise choroid segmentation in OCT images. Building upon RETFound—a foundation model pre-trained on 1.6 million retinal images—RFA-U-Net retains its encoder to leverage rich hierarchical feature representations while addressing its general-purpose segmentation limitations through three key innovations: (1) attention gates to dynamically emphasize choroid-specific features, (2) feature fusion strategies to maintain contextual integrity during decoding, and (3) standard U-Net up-convolutions to ensure accurate boundary reconstruction. Extensive experiments demonstrate that RFA-U-Net outperforms pre-trained CNN-U-Net variants and state-of-the-art (SOTA) choroidal segmentation models. Specifically, it achieves a Dice score of 95.04 ± 0.25% and a Jaccard index of 90.59 ± 0.30% on an independent test set, highlighting its superior accuracy and robustness, particularly in challenging clinical scenarios with variable OCT image quality. These findings underscore the potential of combining foundation models and attention mechanisms to advance precision and reliability in ophthalmic diagnostics, paving the way for scalable and efficient deployment in clinical and community-based screening settings.

## I. INTRODUCTION

The choroid is a dense, vascular layer located between the retina and the sclera, forming the posterior segment of the uvea. It plays a crucial role in ocular health by providing oxygen and nutrients to the outer retina, regulating retinal temperature, and secreting growth factors [1]. Structurally, the choroid is bounded by Bruch’s Membrane (BM) on the upper side and the Choroid-Scleral Interface (CSI) on the lower side. The distance between these two boundaries defines the choroidal thickness, a key biomarker for various ocular diseases such as high myopia, polypoidal choroidal vasculopathy (PCV), age-related macular degeneration (AMD), and diabetic macular edema (DME) [2, 3, 4, 5]. Changes in choroidal thickness have been linked to the progression of these conditions, making choroid segmentation and thickness analysis critical for both clinical and research applications.

Optical Coherence Tomography (OCT) is a high-resolution, noninvasive imaging technique that provides cross-sectional images of the retina and choroid, facilitating the evaluation of structural changes linked to ocular diseases [6]. By performing both qualitative and quantitative analyses of OCT images, clinicians can track disease progression and tailor treatment plans accordingly [7, 8, 9, 10]. To enhance choroidal imaging, various OCT advancements have been introduced, including spectral-domain OCT (SD-OCT) [11], enhanced depth imaging OCT (EDI-OCT) [12], and swept-source OCT (SS-OCT) [13]. While EDI-OCT and SS-OCT are primarily employed for detailed choroidal visualization, SD-OCT is also capable of capturing clear choroidal images, particularly in high myopia cases where the eyeball wall is significantly thinned.

Choroid segmentation in OCT imaging is essential for diagnosing and monitoring choroid-related diseases such as choroiditis, choroidal neovascularization, and central serous chorioretinopathy. Accurate segmentation allows clinicians to measure thickness variations and track disease progression over time. Despite its significance, automated choroid segmentation remains a challenge due to the low contrast between the choroid and sclera, complex vascular textures, and the impact of pathological deformations. Manual segmentation is time-consuming and subjective, whereas existing automatic methods often struggle with anatomical variations and algorithm parameter dependencies [14, 15]. These limitations highlight the need for robust, automated segmentation techniques that can enhance clinical applicability. Prior to the rise of deep learning techniques, choroid segmentation methods were typically categorized into traditional image processing methods and classical machine learning (ML) approaches. Early segmentation approaches relied on graph-based techniques, such as graph search and gradient-based methods [14, 16]. These methods often required manual parameter tuning, resulting in low efficiency and limited clinical application. With the advancement of deep learning (DL), Convolutional Neural Networks (CNNs) have been widely used to improve choroid segmentation accuracy and efficiency [17, 18, 19]. These models leverage end-to-end training to process raw OCT images and generate segmentation results without manual intervention [20, 21]. BEM [22] is one of these methods that introduces a fully automated framework to address the challenge of unclear choroid-sclera boundaries. The authors design a novel Boundary Enhancement Module (BEM), which enhances boundary features through multi-branch processing across feature, channel, and spatial dimensions. By integrating soft boundary point maps and a boundary perceptual loss, the method leverages expert knowledge to improve segmentation accuracy. Evaluated on three datasets, the approach enables detailed 2D and 3D morphological analysis to support studies on high myopia. Following recent efforts to improve automated choroid segmentation, DeepGPET [23] is a DL method that translates the clinically validated GPET approach into a fully automatic system. By fine-tuning a U-Net with a MobileNetV3 backbone, DeepGPET achieves high segmentation accuracy and significantly reduces processing time. It requires no manual intervention, making it well-suited for large-scale datasets and clinical use, particularly in systemic disease studies. In [24], authors introduced PGKD-Net, a two-stage network combining prior knowledge with feature diffusion to handle the low contrast and structural complexity in OCT images. It includes a coarse segmentation stage (PG-Net) guided by a prior mask below the BM layer and a refinement stage (KD-Net) that leverages advanced context aggregation (MSCA) and feature fusion (MLFF) modules. By integrating global contextual cues and cascading multi-level features, PGKD-Net enhances performance across multiple datasets. In response to the limited penetration depth of conventional SD-OCT in visualizing the choroid, in [25], authors present a generative DL model that synthetically enhances SD-OCT scans to approximate the quality of more expensive SS-OCT imaging. Trained on a large paired dataset, the model learns deep anatomical features to generate realistic choroidal structures, enabling accurate extraction of thickness, area, volume, and vascularity metrics. With high correlation to SS-OCT measurements and indistinguishable image quality as judged by experts, this approach expands the diagnostic utility of widely accessible SD-OCT devices in both research and clinical practice. Recent advances have notably improved the precision of choroid segmentation, yet limitations in generalization, boundary clarity, and data efficiency still remain, highlighting the need for further innovation in this field. Recent advances in AI have led to the development of foundation models, a new class of models trained on large, diverse datasets that can be adapted to multiple downstream tasks, including choroid segmentation. Unlike traditional AI models designed for single tasks [26], these models achieve state-of-the-art performance across various problems while demonstrating improved robustness and generalizability [27, 28]. Although still in early stages, their versatility marks a significant shift from earlier task-specific approaches. VisionFM [29] is a foundation model pre-trained on 3.4 million ophthalmic images from diverse diseases, modalities, and demographics, enabling expert-level performance in tasks like disease diagnosis, segmentation, and biomarker prediction. Evaluated on large-scale benchmarks (23 public and 5 private datasets for diagnosis; 23 public and 2 private datasets for segmentation), it outperformed both baseline DL models and ophthalmologists with basic-to-intermediate experience in diagnosing 12 common eye diseases. The model also demonstrated strong generalizability to unseen modalities (e.g., achieving 0.935 AUC in diabetic retinopathy grading on OCTA images despite no prior exposure to OCTA during training). To address data scarcity, VisionFM incorporated synthetic ophthalmic images, validated via visual Turing tests, which enhanced its performance on downstream tasks. While VisionFM relies on supervised pretraining and synthetic data, another foundation model named RETFound [30] emphasizes self-supervision and real-world unlabeled images. This model was trained on 1.6 million unlabeled retinal images (904,170 CFPs and 736,442 OCTs) to address the limitations of task-specific AI models in ophthalmology. By leveraging masked autoencoder-based pretraining, first on natural images (ImageNet-1k) and then on retinal datasets (MEH-MIDAS, EyePACS), RETFound achieves superior label efficiency and generalizability across diverse tasks. It outperforms comparison models (SL-ImageNet, SSL-ImageNet, SSL-Retinal) in diagnosing ocular diseases, predicting progression, and forecasting systemic conditions (e.g., heart failure, Parkinson’s disease) using fewer labeled data.

Building on RETFound’s success in clinical settings, another study demonstrates its real-world utility by developing a RETFound-enhanced DL model for community-based eye disease screening [31]. Trained on challenging real-world images from Shanghai community screenings, the model achieved 15% higher sensitivity and specificity compared to commercial models while showing superior generalization versus traditional AI approaches. Decision curve analysis further confirmed its higher net benefit in both urban and rural Chinese populations, addressing critical resource constraints in low- and middle-income countries. The results validate RET-Found’s adaptability to non-ideal imaging conditions and underscore its potential for scalable public health implementation. While RETFound provides robust feature extraction capabilities, pre-trained on CFPs and OCT scans for diverse ophthalmic tasks, its general-purpose training limits performance for specialized choroidal segmentation. To address this gap, we propose RFA-U-Net, a novel architecture that strategically adapts RETFound for precise choroid layer delineation in OCT images. The model retains RETFound’s encoder to leverage its rich hierarchical representations of retinal structures, while addressing its segmentation limitations through three key innovations: First, attention gates in the decoder dynamically weight choroid-specific features from the encoder layers. Second, these refined features are concatenated with upsampled decoder outputs to maintain contextual information. Third, standard U-Net up-convolutions ensure precise spatial reconstruction of choroid boundaries. This approach uniquely combines RETFound’s broad ophthalmic knowledge with task-specific optimizations, achieving superior performance without computationally expensive retraining. Our solution demonstrates particular effectiveness in challenging clinical scenarios involving variable OCT image quality or ambiguous choroid boundaries, where conventional methods often fail.

The rest of this paper is organized as follows: Section II details the proposed RFA-U-Net framework, including its architecture and refinement techniques. Section III describes the experimental setup, presents the results, and analyzes the contributions of this study. Section IV discusses the findings and concludes the study by summarizing key insights, highlighting the impact of RFA-U-Net, and outlining potential directions for future research. Finally, Sections V and VI provide information on data and code availability.

### II. MATERIALS AND METHODS

### A. Dataset and Ethical Approval

In this study, we used an initial dataset of 966 OCT images, each paired with a manually annotated ground truth mask delineating the choroid layer. The annotations were performed by a research assistant with over one year of experience in image analysis and subsequently reviewed by an ophthalmologist specializing in uveitis. Initially, the full dataset was divided into a training, validation, and test set, with 80% (773 images) allocated for training and validation, and 20% (193 images) for testing. The training/validation set was further partitioned into five folds using 5-fold cross-validation to assess the generalization performance of the models. Each fold was used as a validation set once, while the remaining four folds served as training data, allowing for a thorough evaluation of the model across different subsets of the data. This cross-validation approach ensured that all images in the training/validation set were used for both training and validation at some point, reducing the risk of overfitting. The test set, comprising 20% of the dataset, was kept completely independent and was only used to evaluate the final performance of the trained models. The random seed set throughout the process ensured the consistency of the train-validation-test split and the reproducibility of the results.

### B. Image Preprocessing

A standardized image preprocessing pipeline was applied uniformly across all models to ensure consistency in input data handling. The preprocessing steps included resizing, where all OCT images were scaled to 224 *×* 224 pixels to align with the input requirements of the neural network architectures. Normalization was performed by scaling pixel intensity values to the range [0, 1], facilitating faster convergence during training. Additionally, data augmentation techniques were employed exclusively on the training dataset to enhance the model’s generalization capabilities. The data augmentation was implemented using a custom JointTransform class, ensuring that both the images and their corresponding masks underwent identical transformations. This approach maintains the alignment between images and masks, which is crucial for accurate segmentation. Finally, the masks were one-hot encoded to align with the output dimensions of the models.

### C. Choroidal Layer Segmentation

After completing the preprocessing of the OCT images, we utilized several models to perform choroidal layer segmentation. The subsequent sections detail the model architectures, which include pre-trained convolutional neural networks (CNN-U-Net) based on architectures such as AlexNet, Inception, VGG19, VGG16, ResNet101, and ResNet50 combined with U-Net. Additionally, we explore more recent state-of-the-art models tailored for choroidal layer segmentation, including U-Net-BEM, DeepGPET, PGKD-Net, and finally, we introduce our proposed model, RFA-U-Net.

1) ***Pre-trained CNN-U-Net***: In this section, we employed pre-trained CNN-U-Net-based models that integrate well-established DL architectures such as AlexNet, Inception, VGG, and ResNet with the U-Net framework. These models leverage the strengths of both pre-trained CNNs and U-Net’s specialized segmentation architecture, making them highly effective for choroid segmentation.

To incorporate pre-trained models, we utilized transfer learning, a widely adopted technique that allows models to adapt to new tasks by leveraging knowledge gained from prior training on large-scale datasets. As comprehensively surveyed by Zhuang et al. [32], transfer learning enables neural networks to generalize effectively to new domains, even when labeled training data is limited. By integrating transfer learning with the U-Net architecture, our models benefit from the feature extraction capabilities of pre-trained CNNs while preserving U-Net’s ability to capture fine-grained spatial details essential for segmentation tasks. This combination enhances both accuracy and efficiency, enabling the models to effectively delineate the choroidal layer in OCT images. A detailed summary of each utilized model’s architecture, including its CNN backbone, is provided in Table I.

**TABLE I.** Summary of Model Architectures for Pre-trained CNN-U-Net-Based Approaches.

| Model Name | Encoder | Bottleneck | Expanding Path | Output Layer | Notes |
| --- | --- | --- | --- | --- | --- |
| AlexNet-U-Net | Pre-trained **AlexNet** | Max Pooling, Conv layer 512 channels, Batch Norm. | Transposed convolutions for upsampling, skip connections with feature concatenation | $1 \times 1$ Conv, 2 classes, Bilinear | Utilizes first five blocks of AlexNet |
| Inception-U-Net | Pre-trained **Inception** | Inception module with 512 channels, Batch Norm. | Mirrors encoder with upsampling, skip connections, feature concatenation | $1 \times 1$ Conv, 2 classes, Bilinear | Combines multi-scale feature extraction by Inception models |
| VGG19-U-Net | Pre-trained **VGG19** | Convolutional layers refining 512-channel, Batch Norm. | Transposed convolutions for upsampling, skip connections, feature concatenation | $1 \times 1$ Conv, 2 classes, Bilinear | Uses first 29 layers of VGG19, excludes final max-pool |
| VGG16-U-Net | Pre-trained **VGG16** | Feature maps refined by additional convolutional layers, Batch Norm. | Transposed convolutions for upsampling, skip connections, feature concatenation | $1 \times 1$ Conv, 2 classes, Bilinear | Based on VGG16's five convolutional blocks |
| ResNet101-U-Net | Pre-trained **ResNet101** | Deep residual features processed with upsampling and concatenation | Bilinear upsampling, skip connections with feature concatenation, Dropout | $1 \times 1$ Conv, 2 classes, Bilinear | Utilizes ResNet101's residual blocks |
| ResNet50-U-Net | Pre-trained **ResNet50** | Deep residual features processed with upsampling and concatenation | Bilinear upsampling, skip connections with feature concatenation, Dropout | $1 \times 1$ Conv, 2 classes, Bilinear | Utilizes ResNet50's residual blocks |

2) ***State-of-the-Art Models for Choroidal Layer Segmentation***: In addition to pre-trained CNN-U-Net variants, we have examined three cutting-edge choroid segmentation models for comparative benchmarking.

#### U-Net-BEM

Wu et al. [22] present a boundary-focused encoder-decoder network for choroid segmentation, termed U-Net-BEM. The Boundary Enhancement Module (BEM) operates at each encoder-decoder level, emphasizing choroid–sclera boundaries through: Multi-scale feature extraction, Channel-level boundary accentuation, and an optimized activation function. By incorporating expert key point maps, U-Net-BEM out-performs standard U-Net variants in delineating difficult choroidal interfaces.

#### DeepGPET

Muller et al. [23] address the choroidal binarization task, separating vascular from stromal tissue, using fully convolutional networks (e.g., U-Net and SegNet). Findings underscore the effectiveness of deep learning for quantifying choroidal vascularity, an important metric in both clinical and research settings.

#### PGKD-Net

Wang et al. [24] propose a two-stage approach combining a Prior-mask Guided Network (PG-Net) for coarse segmentation with a Knowledge Diffusive Network (KD-Net) for refined results. Key modules include: Multi-Scale Context Aggregation (MSCA) and Multi-Level Feature Fusion (MLFF). MSCA captures non-local dependencies to improve global context understanding, and MLFF integrates coarse outputs into refined feature representations for robust segmentation. Their open-source implementation demonstrates state-of-the-art accuracy, highlighting the importance of multi-level feature processing in challenging boundary conditions.

3) ***RFA-U-Net***: In developing U-Net-based models for segmenting the choroid layer in OCT images, integrating pre-trained CNNs as encoders leverages transfer learning to enhance feature extraction capabilities [33, 34]. The U-Net architecture, introduced by Ronneberger et al., is renowned for its effectiveness in biomedical image segmentation [35]. The RFA-U-Net model introduced in this study integrates the RETFound foundation model as its encoder within the traditional U-Net architecture [35]. By incorporating the RETFound encoder, RFA-U-Net leverages pre-trained knowledge from a vast retinal image repository, thereby improving feature extraction and segmentation accuracy. RETFound has been validated in multiple disease detection tasks, demonstrating its versatility and robustness. Additionally, RETFound can be efficiently adapted to customized tasks, making it an ideal choice for the specific requirements of choroid layer segmentation in OCT images. The proposed architecture comprises the following components.

#### RETFound Encoder (Contracting Path)

RET-Found is a SOTA foundation model for retinal imaging, pre-trained on 1.6 million retinal images using self-supervised learning methodologies based on Masked Autoencoders (MAE). This extensive pre-training enables RETFound to capture intricate patterns and features inherent in retinal images, facilitating improved generalization across various disease detection tasks. The encoder extracts hierarchical features at multiple resolutions, which are crucial for precise segmentation [30].

#### Attention Gates

Attention gates play a crucial role in neural network architectures, especially for medical image segmentation. They work by selectively highlighting important features while filtering out less relevant information, which helps the model concentrate on the critical regions necessary for accurate segmentation. This is achieved by computing attention coefficients using features from both the encoder and decoder, enabling the network to dynamically emphasize significant spatial areas. Incorporating attention gates into architectures such as U-Net has demonstrated improved segmentation accuracy by allowing the model to better focus on meaningful regions and disregard unnecessary details [36, 37].

#### Bottleneck

Serving as the bridge between the encoder and decoder, the bottleneck consists of convolutional layers that further process the high-level features extracted by the encoder. This component refines the feature representations before they are upsampled in the decoder, ensuring that the most salient information is retained for subsequent reconstruction.

#### Decoder (Expanding Path)

The decoder reconstructs the spatial resolution of the feature maps through a series of upsampling operations. Each decoder block includes an upsampling layer, followed by a convolutional layer that reduces the number of feature channels. The attention gates facilitate the integration of skip connections from the encoder by weighting the encoder features based on their relevance. This process ensures that fine-grained details are effectively merged with the upsampled features, enhancing the precision of the segmentation output.

#### Output Layer

The final layer is a 1×1 convolutional layer that maps the decoder’s output to the desired number of segmentation classes. A Softmax activation function is applied to generate probability maps corresponding to each class, producing segmentation masks with the shape of (2,224,224).

Fig. 1 illustrates the RFA-U-Net architecture, highlighting its advanced design tailored for precise feature extraction and segmentation.

**Fig. 1.**
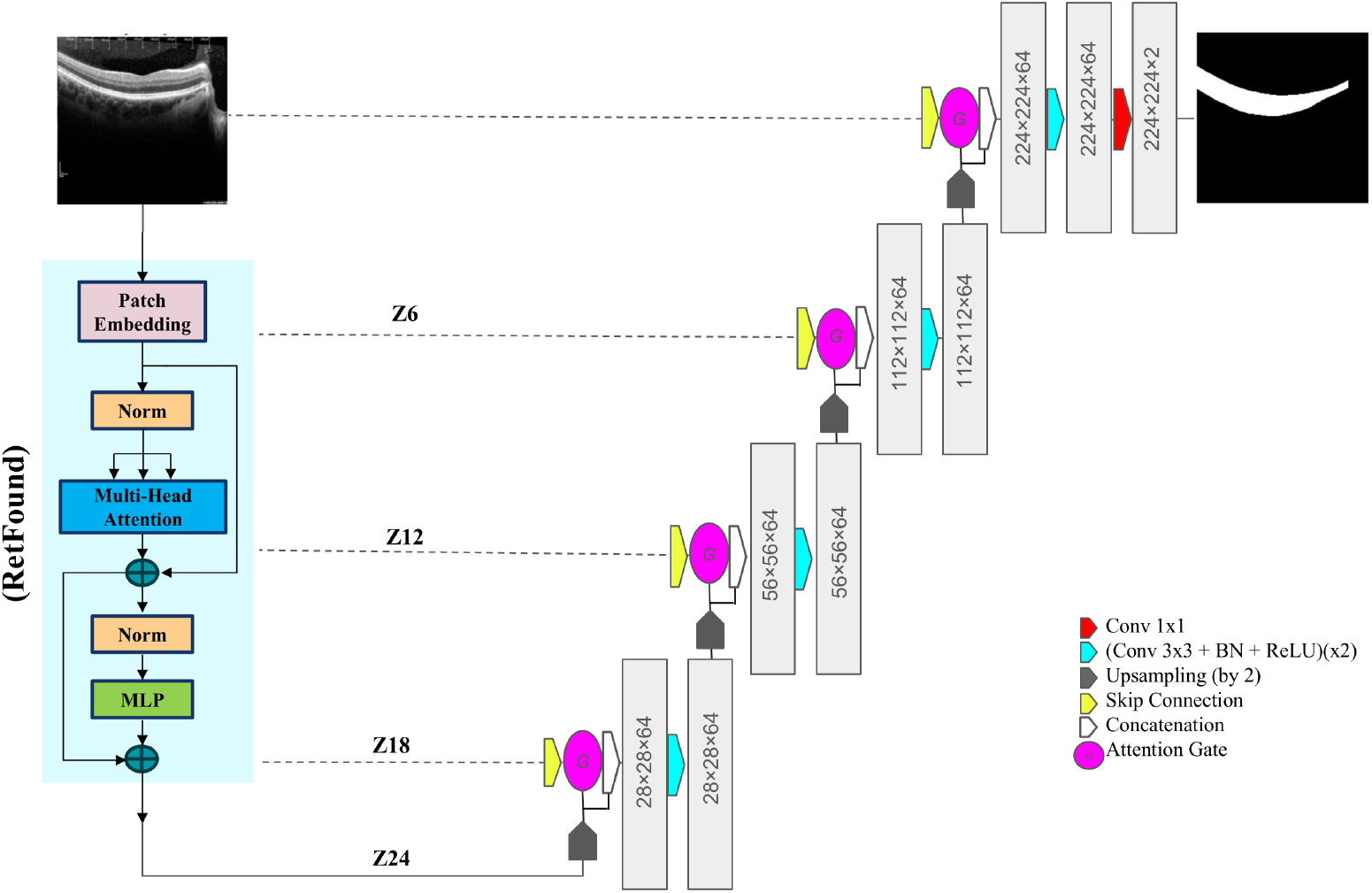
Detailed Overview of the RFA-U-Net Architecture for Enhanced Feature Extraction and Segmentation.

### D. Training Configuration

All models were trained under consistent configurations to ensure a fair comparison, with particular settings tailored for the RFA-U-Net model. Tables II and III present the optimized hyperparameters for two distinct groups: CNN-U-Net-based models and SOTA models.

**TABLE II.** Tuned Hyperparameters for Pre-trained CNN-U-Net Models and Proposed Model.

| Hyperparameter | AlexNet-U-Net | Inception-U-Net | VGG19-U-Net | VGG16-U-Net | ResNet101-U-Net | ResNet50-U-Net | RFA-U-Net |
| --- | --- | --- | --- | --- | --- | --- | --- |
| Learning Rate | 1e-4 |  |  |  |  |  |  |
| Batch Size | 8 |  |  |  |  |  |  |
| Number of Epochs | 150 |  |  |  |  |  |  |
| Optimizer | Adam |  |  |  |  |  |  |
| Scheduler | ReduceLROnPlateau |  |  |  |  |  |  |
| Pretrained Weights | ImageNet | ImageNet | ImageNet | ImageNet | ImageNet | ImageNet | MAE (Self-Supervised on 1.6 million retinal images) |
| Early Stopping Patience | 10 | 10 | 10 | 10 | 10 | 10 | 8 |
| Dropout Rate | 0.5 | 0.5 | 0.5 | 0.5 | 0.5 | 0.5 | Not used |
| Number of Layers | 16 | 16 | 19 | 16 | 101 | 50 | 24 |
| Activation Function | SoftMax |  |  |  |  |  |  |
| Batch Normalization | Yes |  |  |  |  |  |  |
| Class Weights (for Loss) | [1.0, 2.0] |  |  |  |  |  |  |
| Input Size | 224x224 |  |  |  |  |  |  |
| Output Size | 224x224 |  |  |  |  |  |  |
| Frozen Layers | 3 | 0 | 1 | 2 | 3 | 3 | 21 |
| Loss Function | Cross-Entropy |  |  |  |  |  | Tversky Loss |

**TABLE III.**
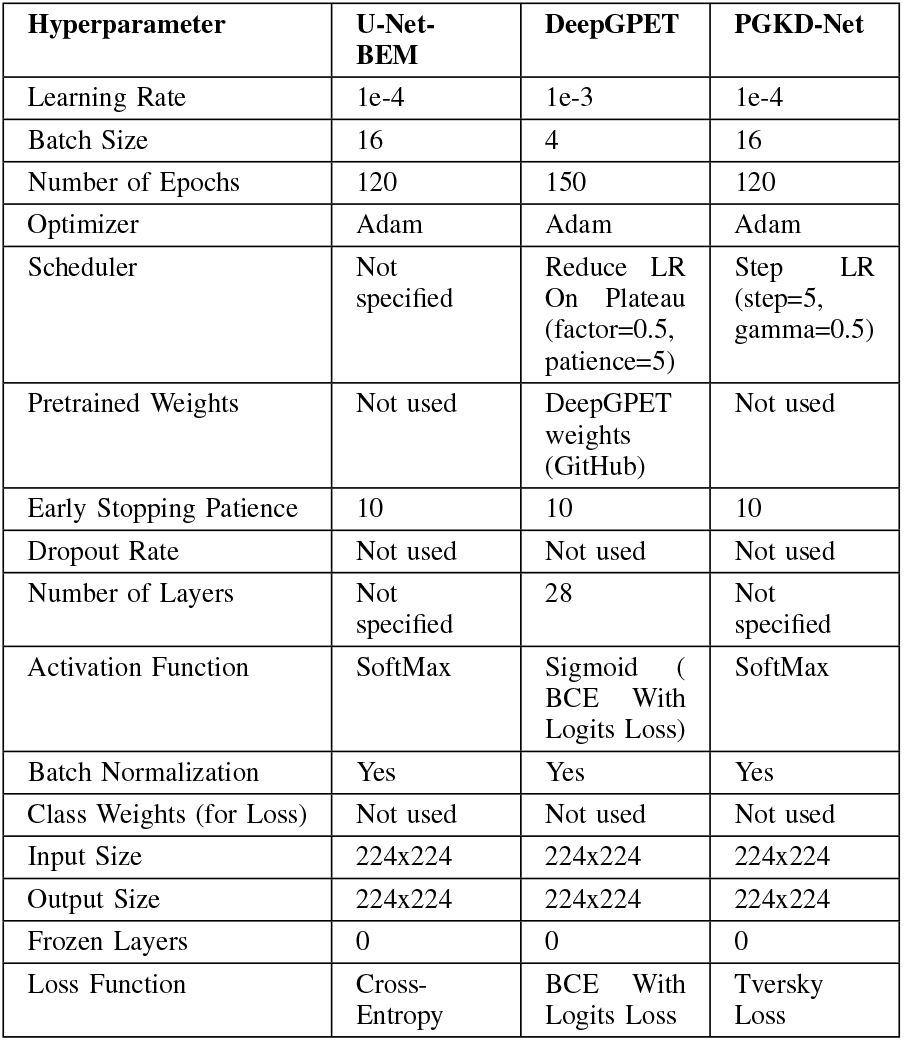
Tuned Hyperparameters for SOTA Models.

| Hyperparameter | U-Net-BEM | DeepGPET | PGKD-Net |
| --- | --- | --- | --- |
| Learning Rate | 1e-4 | 1e-3 | 1e-4 |
| Batch Size | 16 | 4 | 16 |
| Number of Epochs | 120 | 150 | 120 |
| Optimizer | Adam | Adam | Adam |
| Scheduler | Not specified | Reduce LR On Plateau (factor=0.5, patience=5) | Step LR (step=5, gamma=0.5) |
| Pretrained Weights | Not used | DeepGPET weights (GitHub) | Not used |
| Early Stopping Patience | 10 | 10 | 10 |
| Dropout Rate | Not used | Not used | Not used |
| Number of Layers | Not specified | 28 | Not specified |
| Activation Function | SoftMax | Sigmoid (BCE With Logits Loss) | SoftMax |
| Batch Normalization | Yes | Yes | Yes |
| Class Weights (for Loss) | Not used | Not used | Not used |
| Input Size | 224x224 | 224x224 | 224x224 |
| Output Size | 224x224 | 224x224 | 224x224 |
| Frozen Layers | 0 | 0 | 0 |
| Loss Function | Cross-Entropy | BCE With Logits Loss | Tversky Loss |

### E. Evaluation Metrics

The following evaluation metrics were consistently applied across all models to exclusively assess the segmentation performance of the choroidal layer.

Dice Coefficient: Measures the overlap between the predicted segmentation of the choroidal layer and the ground truth.

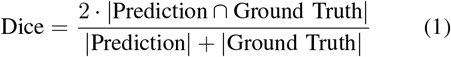

Jaccard Index (Intersection over Union): Evaluates the similarity between predicted and true masks.

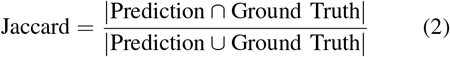

Boundary Errors: Upper and Lower Bound Signed Errors (UBSE, LBSE) Calculate the average signed distance between the predicted and true boundaries of the choroidal layer. Upper and Lower Bound Unsigned Errors (UBUE, LBUE) Calculate the average absolute distance between the predicted and true boundaries of the choroidal layer. It is worth noting that in this study, the Dice and Jaccard coefficients were calculated exclusively within the region of interest (ROI) rather than the entire image. These metrics were consistently applied across all models to assess the segmentation performance of the choroidal layer. In segmentation tasks, focusing on the ROI, specifically the choroidal layer in this study, is crucial to ensure accurate and meaningful results. Including irrelevant areas, such as background regions, in the calculation can distort the evaluation by inflating performance scores. This occurs because extensive background regions may dominate the assessment, masking segmentation errors within the choroidal layer and leading to an overestimation of model performance.

## III. RESULTS

In this study, a method based on the RETFound was introduced for choroid layer segmentation. By retaining RETFound’s encoder and integrating attention-gated decoding, contextual feature fusion, and standard U-Net up-convolutions, RFA-U-Net achieves precise spatial delineation of choroid boundaries. This design leverages pretrained retinal representations while effectively addressing task-specific challenges.

The proposed model, along with baseline models, was rigorously evaluated on an independent test set consisting of 193 OCT images to comprehensively assess its generalization capabilities. All experiments were conducted using an NVIDIA Tesla A100 GPU with approximately 16 GB of dedicated GPU memory on Google Colab. The implementation was carried out in Python, version 3.11.11. The results demonstrate the superior performance and robustness of our model, as detailed in the following subsections.

### A. Performance of the Proposed Model (RFA-U-Net)

The proposed model demonstrated exceptional performance across multiple quantitative metrics. It achieved a Dice Score of 95.04 ± 0.25% and Jaccard index of 90.59 ± 0.30%, underscoring its remarkable ability to accurately capture the overlap between predicted and ground truth regions. For boundary measurements, the model achieved an Upper Boundary Signed Error (UBSE) of −0.89 ± 0.10 µm and an Upper Boundary Unsigned Error (UBUE) of 6.04 ± 0.20 µm, as well as a Lower Boundary Signed Error (LBSE) of 1.05 ± 0.15 µm and a Lower Boundary Unsigned Error (LBUE) of 21.4 ± 1.00 µm. These results highlight the model’s precision and reliability in delineating the choroidal layer in OCT images. Fig. 2 provides a visual representation of the model’s performance, showcasing its effectiveness in segmentation tasks. The figure illustrates the predicted and ground truth upper and lower boundaries of the choroidal layer across several test B-scan samples, further demonstrating the strong performance of the RFA-U-Net model.

**Fig. 2.**
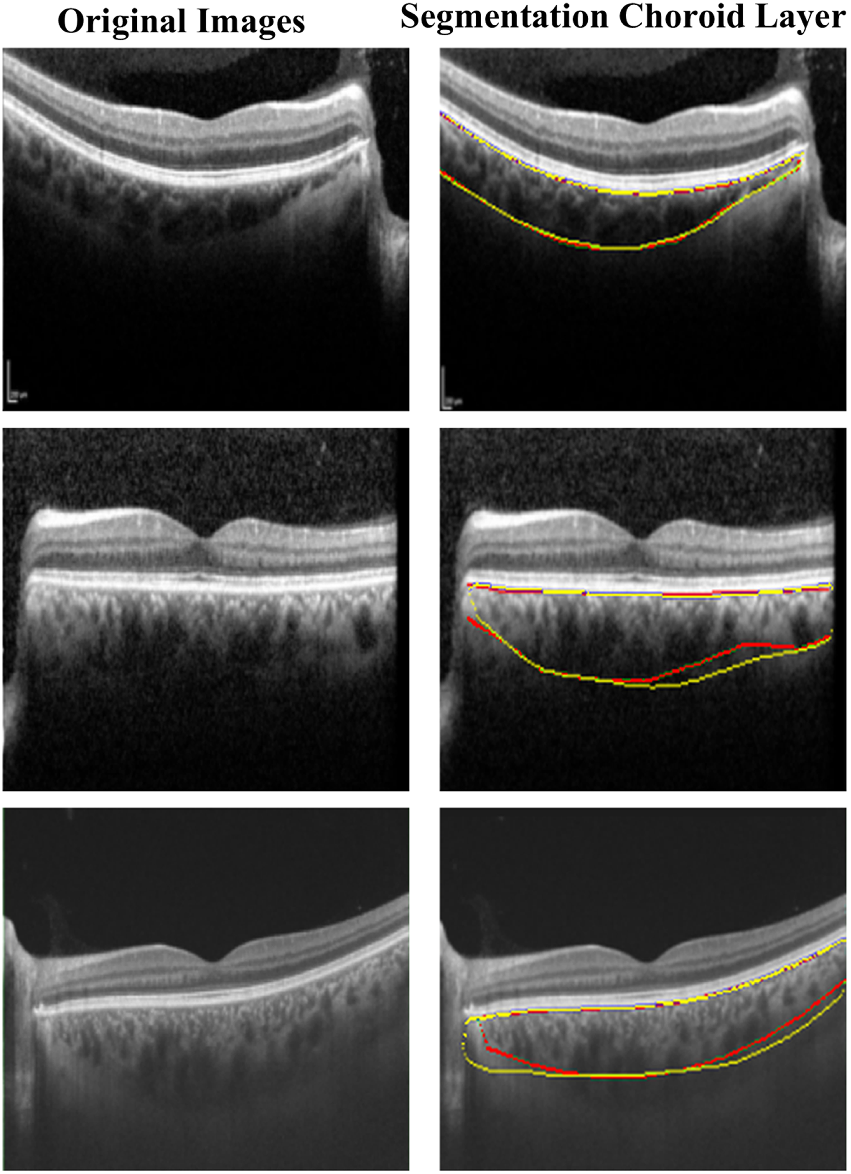
Segmentation of the choroid layer using RFA-U-Net. The red boundary represents the ground truth, and the yellow boundary denotes the predicted segmentation. The left column displays the corresponding original images, while the right column presents the segmented results.

### B. Comparison of the RFA-U-Net with Other Models

To thoroughly evaluate the performance of the proposed model, a comprehensive comparison was conducted against several pre-trained CNN-U-Net models and recent state-of-the-art (SOTA) methods specifically designed for choroidal segmentation. This analysis aimed to assess the accuracy, robustness, and overall effectiveness of RFA-U-Net relative to established benchmarks in the field.

As shown in Tables IV and V, the proposed RFA-U-Net consistently achieved superior results across both overlap-based metrics (Dice and Jaccard) and boundary-based error metrics (UBSE, UBUE, LBSE, LBUE). In comparison with CNN-U-Net baselines, including AlexNet-U-Net, VGG19-U-Net, and ResNet50-U-Net, RFA-U-Net achieved the highest Dice score and Jaccard index, while simultaneously minimizing boundary errors. In particular, it exhibited the lowest LBUE, a critical metric for accurately delineating the choroid layer, highlighting its robust spatial precision over traditional CNN-based architectures.

**TABLE IV.** Comprehensive Evaluation of Test Set Results Across Pre-trained CNN-U-Net models and the proposed model. The best results are highlighted in Bold, with values presented in the format Mean ± std.

| Model | Dice (%) | Jaccard (%) | UBSE ($\mu\text{m}$) | UBUE ($\mu\text{m}$) | LBSE ($\mu\text{m}$) | LBUE ($\mu\text{m}$) |
| --- | --- | --- | --- | --- | --- | --- |
| AlexNet-U-Net | 94.11 ± 0.30 | 89.03 ± 0.35 | -2.36 ± 0.20 | 7.62 ± 0.25 | 4.5 ± 0.20 | 27.51 ± 1.20 |
| Inception-U-Net | 93.47 ± 0.28 | 88.31 ± 0.33 | -0.27 ± 0.10 | 5.6 ± 0.18 | 2.15 ± 0.15 | 28.19 ± 1.10 |
| VGG19-U-Net | 93.34 ± 0.25 | 87.33 ± 0.30 | -1.56 ± 0.15 | 5.11 ± 0.22 | -2.46 ± 0.18 | 28.3 ± 1.00 |
| VGG16-U-Net | 92.95 ± 0.30 | 86.91 ± 0.35 | 3.69 ± 0.20 | 7.13 ± 0.25 | 4.3 ± 0.20 | 29.61 ± 1.20 |
| ResNet101-U-Net | 92.67 ± 0.28 | 87.06 ± 0.32 | -1.42 ± 0.15 | 5.35 ± 0.22 | 1.44 ± 0.15 | 28.52 ± 1.10 |
| ResNet50-U-Net | 92.06 ± 0.30 | 85.93 ± 0.35 | 0.09 ± 0.12 | 6.46 ± 0.23 | 2.59 ± 0.18 | 31.2 ± 1.20 |
| RFA-U-Net | **95.04 ± 0.25** | **90.59 ± 0.30** | -0.89 ± 0.10 | 6.04 ± 0.20 | 1.05 ± 0.15 | **21.4 ± 1.00** |

**TABLE V.**
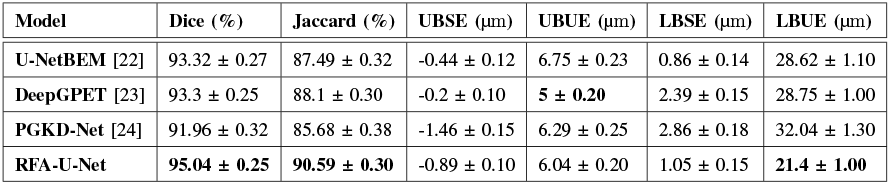
Comprehensive Evaluation of Test Set Results Across State Of Art models and the proposed model. The best results are highlighted in Bold, with values presented in the format Mean ± std.

| Model | Dice (%) | Jaccard (%) | UBSE ($\mu\text{m}$) | UBUE ($\mu\text{m}$) | LBSE ($\mu\text{m}$) | LBUE ($\mu\text{m}$) |
| --- | --- | --- | --- | --- | --- | --- |
| U-NetBEM [22] | 93.32 ± 0.27 | 87.49 ± 0.32 | -0.44 ± 0.12 | 6.75 ± 0.23 | 0.86 ± 0.14 | 28.62 ± 1.10 |
| DeepGPET [23] | 93.3 ± 0.25 | 88.1 ± 0.30 | -0.2 ± 0.10 | **5 ± 0.20** | 2.39 ± 0.15 | 28.75 ± 1.00 |
| PGKD-Net [24] | 91.96 ± 0.32 | 85.68 ± 0.38 | -1.46 ± 0.15 | 6.29 ± 0.25 | 2.86 ± 0.18 | 32.04 ± 1.30 |
| RFA-U-Net | **95.04 ± 0.25** | **90.59 ± 0.30** | -0.89 ± 0.10 | 6.04 ± 0.20 | 1.05 ± 0.15 | **21.4 ± 1.00** |

To further validate its effectiveness, RFA-U-Net was compared against specialized SOTA methods, including U-NetBEM, DeepGPET, and PGKD-Net (Table V). Despite these methods being explicitly developed for choroidal segmentation, RFA-U-Net outperformed them in most key metrics. While DeepGPET achieved a slightly lower UBUE (5 ± 0.20 µm), RFA-U-Net demonstrated a markedly higher Dice score (approximately 1.7% higher) and a substantially lower LBUE (21.4 ± 1.00 µm versus 28.75 ± 1.00 µm), indicating a better balance between segmentation accuracy and boundary precision, an essential consideration for clinical applications.

These results strongly highlight that RFA-U-Net not only surpasses conventional segmentation baselines but also competes favorably against specialized SOTA methods, offering a robust, accurate, and computationally efficient solution for choroid layer delineation in OCT images. The model’s superiority is further illustrated in the radar chart (Figs. 3 and 4), which visually summarizes the performance of different models across key evaluation metrics, including Dice, Jaccard, and boundary errors (UBUE and LBUE). To maintain a consistent interpretation where larger values indicate better performance, the boundary error metrics were inverted, plotted as 1-UBUE and 1-LBUE. As shown, RFA-U-Net consistently achieves the best overall performance, with higher Dice and Jaccard scores and reduced boundary errors compared to both CNN-U-Net-based models and recent SOTA methods. These results highlight RFA-U-Net’s superior balance between segmentation accuracy and boundary delineation, underscoring its effectiveness for precise choroidal layer segmentation.

**Fig. 3.**
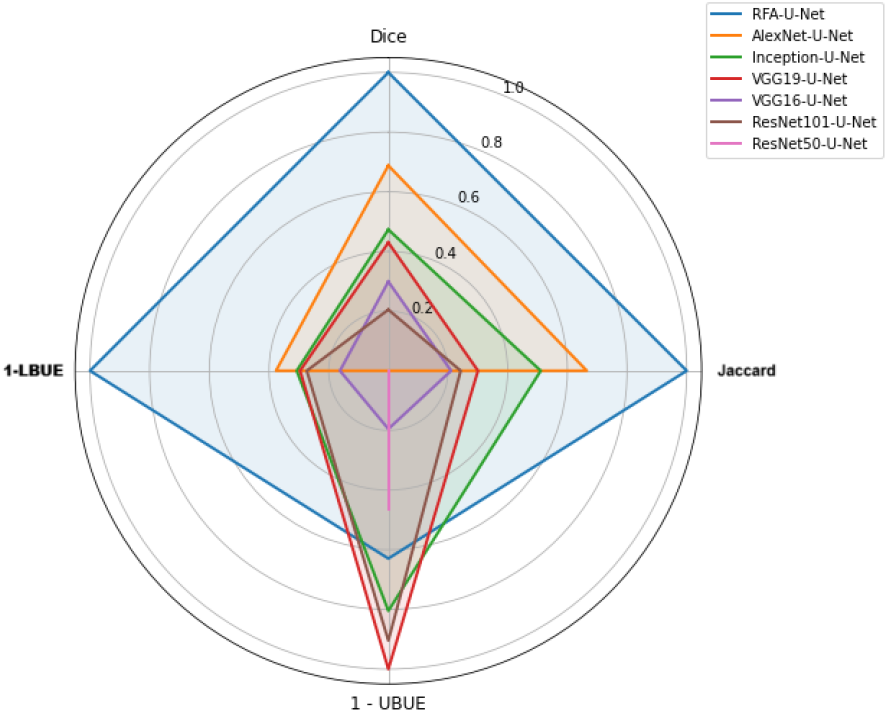
Radar chart illustrating the performance of Pre-trained CNN-U-Net models and the proposed model on the test set.

**Fig. 4.**
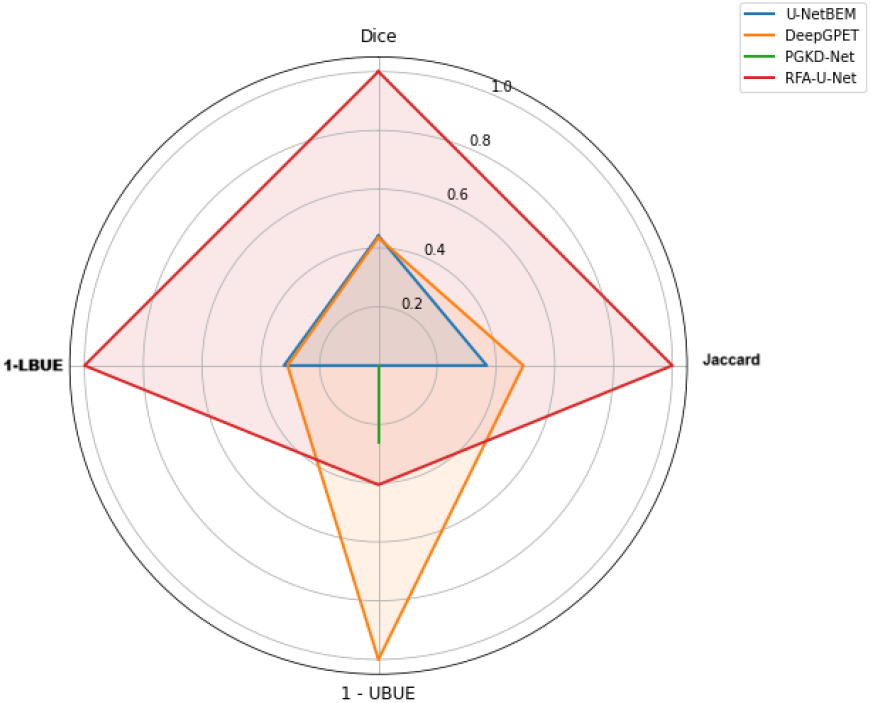
Radar chart illustrating the performance of SOTA models and the proposed model on the test set.

To effectively showcase the performance of the proposed model in comparison to others, several test samples were selected, with the ground truth and the predicted upper and lower boundaries of the choroidal layer highlighted for evaluation. These visual comparisons were performed against pre-trained CNN-U-Net models (Fig. 5) and other SOTA models in the field (Fig. 6). The results clearly underscore the superior ability of the proposed model to accurately delineate the boundaries of the choroidal layer. This advantage is particularly pronounced in regions with complex anatomical variations, where the proposed model consistently outperforms alternative approaches, emphasizing its robustness and precision.

**Fig. 5.**
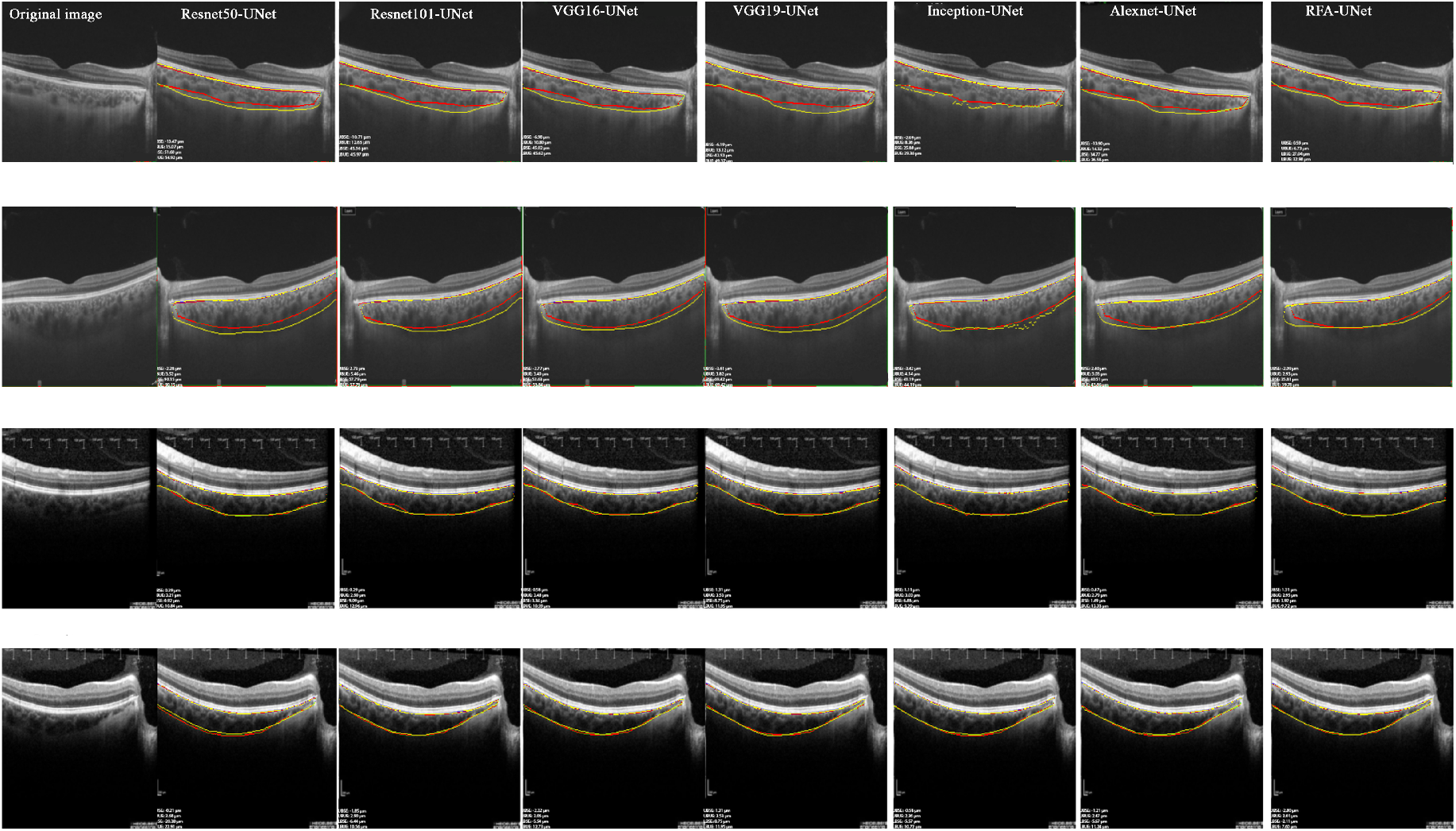
Comparative segmentation performance of RFA-UNet and pre-trained CNN-U-Net models on several test samples. The red boundary represents the ground truth, while the yellow boundary depicts the model’s predictions. Each row represents the results for one test sample, while each column corresponds to the results of one model for that test sample. Metrics including UBSE (Upper Boundary Signed Error), UBUE (Upper Boundary Unsigned Error), LBSE (Lower Boundary Signed Error), and LBUE (Lower Boundary Unsigned Error) are displayed in the lower-left corner of each image.

**Fig. 6.**
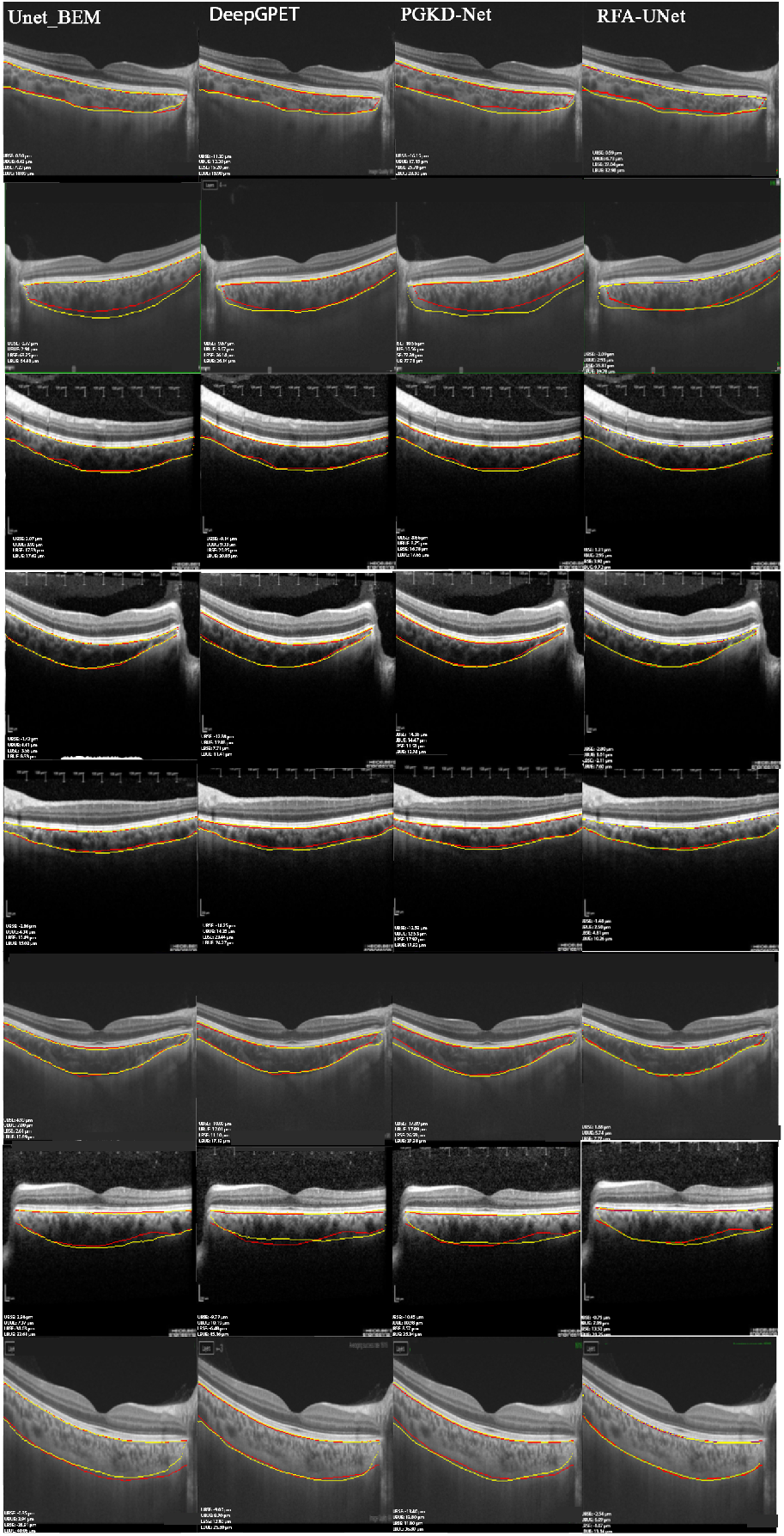
Comparative visualization of segmentation results by RFA-U-Net, and other SOTA models in the field on the selected test samples. Each image displays the segmentation boundaries with the red boundary representing the ground truth and the yellow boundary indicating the predicted boundary by the respective model. Each row represents the results for one test sample, while each column corresponds to the results of one model for that test sample. In the lower-left corner of each image, the following metrics are reported: UBSE (Upper Boundary Signed Error), UBUE (Upper Boundary Unsigned Error), LBSE (Lower Boundary Signed Error), and LBUE (Lower Boundary Unsigned Error).

Figure 7 compares the Dice scores across all models in three categories: The Proposed Model, Pre-trained CNN-U-Net Models, and SOTA Models. As can be seen, the RFA-U-Net model achieves the highest Dice score of 95%, surpassing all other models. The Pre-trained CNN-UNet models show a decline from 94% to 92%, while SOTA models fluctuate between 92% and 93%. This highlights RFA-U-Net’s superior performance in the segmentation task.

**Fig. 7.**
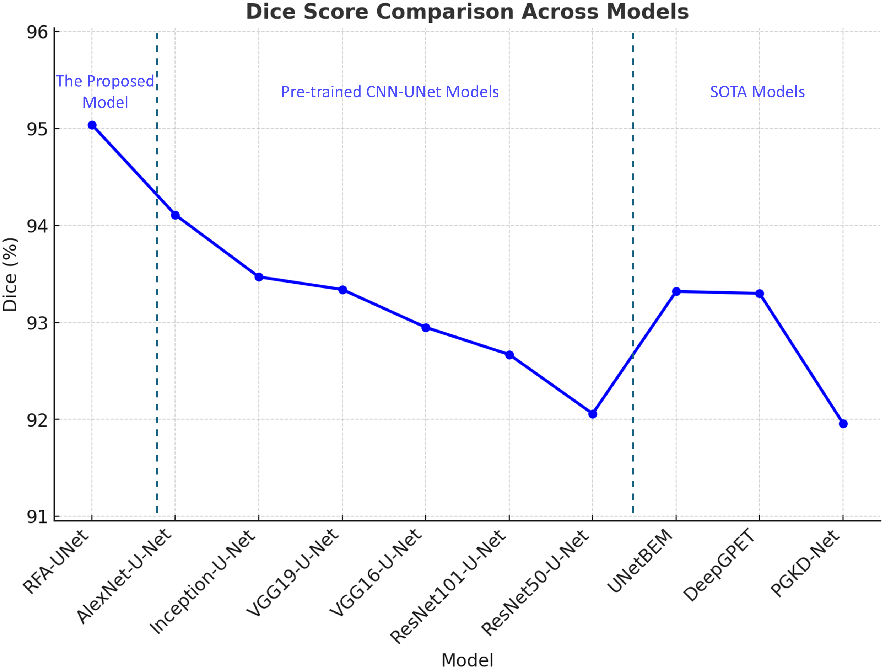
Comparison of Dice Scores Across All Models in Three Categories.

Figure 8 displays a plot comparing upper and lower boundary errors across three categories based on µm. The Proposed Model, RFA-U-Net, exhibits the lowest lower boundary error, indicating strong performance. In contrast, AlexNet-U-Net shows the highest upper boundary error, while PGKD-Net has the highest lower boundary error.

**Fig. 8.**
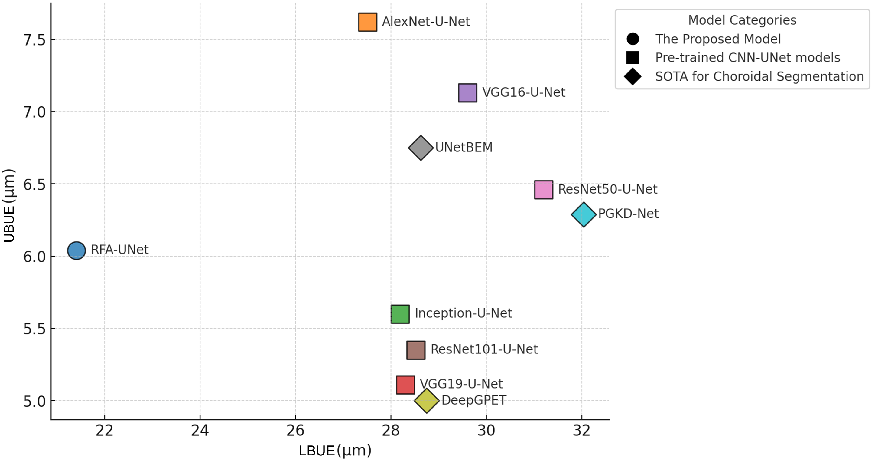
Comparison of Upper and Lower Boundary Errors Across All Models in Three Categories for Choroidal Segmentation.

Figure 9 presents correlation plots comparing ground truth and model-derived measures of mean choroid thickness (left) and choroid area (right) using RFA-U-Net. For choroid thickness, RFA-U-Net achieves a high Pearson’s r of 0.94, with a linear fit of y = 0.87x + 0.03, closely aligning with the ideal line (y = x), indicating strong agreement. Similarly, for choroid area, RFA-U-Net shows a Pearson’s r of 0.95, with a linear fit of y = 0.90x + 0.06, also near the ideal line. These results demonstrate RFA-U-Net’s high accuracy and reliability in choroidal segmentation, with minimal systematic bias.

**Fig. 9.**
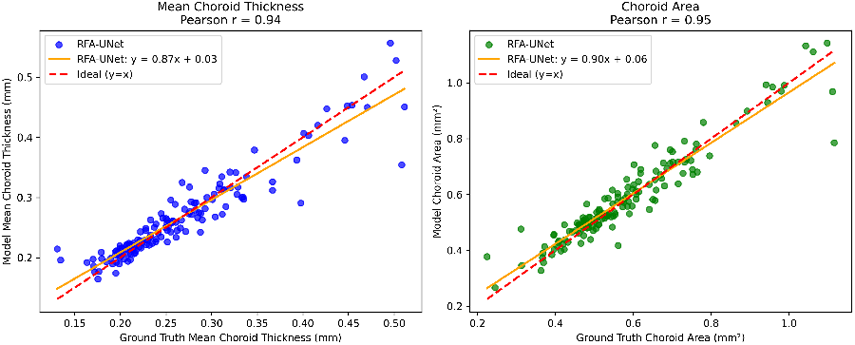
Correlation Plots of Mean Choroid Thickness and Area Using RFA-U-Net.

## IV. DISCUSSION

Accurate segmentation of the choroid layer is essential in both research and clinical settings. In diseases such as central serous chorioretinopathy or polypoidal choroidal vasculopathy, changes in choroid thickness and boundaries are diagnostic biomarkers [38, 39]. Moreover, choroidal microvascular metrics improve discriminative power for diabetic retinopathy when added to retinal parameters [40]. Automated segmentation, can reduce manual workload while providing consistent and reproducible measurements. This capability could significantly enhance early detection, monitoring, and personalized treatment strategies. In this study, we introduced the RFA-U-Net model for choroid segmentation in OCT images, leveraging the RETFound foundation model as the encoder and integrating attention mechanisms in the decoder. This integration adds a unique dimension to the RFA-U-Net model. RETFound’s extensive pre-training on a large retinal image repository ensures robust feature extraction, even with limited labeled data [30, 41]. This positions the RFA-U-Net as a versatile tool for retinal imaging tasks beyond choroid segmentation, including disease classification and multi-layer analysis [42]. Across our dataset of 966 OCT images—partitioned into training, validation, and test subsets—RFA-U-Net demonstrated the highest Dice Score (95.04) and Jaccard Index (90.59), significantly outperforming alternative pre-trained CNN-U-Net-based architectures (e.g., VGG16-U-Net, ResNet101-U-Net) and SOTA methods in segmentation of choroid layer (PGKD-Net, U-NetBEM, and DeepGPET). Furthermore, its lower boundary errors, especially in the critical lower unsigned error, underscore its capability for precise delineation of choroid layer boundaries. Pretrained CNN-U-Net models, such as VGG16-U-Net and ResNet101-U-Net, utilize encoders pre-trained on large-scale classification tasks, leveraging transfer learning to enhance feature extraction. However, they may be less effective in capturing specialized retinal features compared to RET-Found. The attention gates integrated into the RFA-U-Net decoder improve spatial selectivity, surpassing standard skip connections in achieving detailed feature mapping. In this study, the inclusion of attention gates in the RFA-U-Net decoder facilitated better focus on relevant image regions, enhancing segmentation precision [41]. Additionally, the utilization of Tversky loss addressed class imbalances, improving segmentation performance for smaller or irregularly shaped structures [43]. These findings align with prior research highlighting the effectiveness of attention mechanisms and customized loss functions in medical image segmentation [34]. This comparison extends to other SOTA models for choroidal layer segmentation. U-NetBEM enhances the boundary at the transition between the choroid and sclera using Boundary Enhancement Modules (BEM), improving boundary refinement but not effectively compensating for errors in transitional zones. In contrast, RFA-U-Net employs hierarchical encoding and attention gates to refine boundary details at a localized level, effectively mitigating errors in these transition areas. Similarly, while DeepGPET achieves high segmentation accuracy in distinguishing stromal from vascular tissue, its reliance on a predefined Gaussian Process Edge Tracing (GPET) algorithm limits its adaptability to complex boundary variations. On the other hand, RFA-U-Net, designed for precise pixel-wise segmentation of choroidal structures, demonstrates superior performance in boundary-sensitive evaluations compared to alternative methods. PGKD-Net adopts a two-stage segmentation approach—Prior-mask Guided Network (PG-Net) for coarse segmentation and Knowledge Diffusive Network (KD-Net) for refinement, achieving high segmentation accuracy. However, its dependency on prior-mask generation reduces its generalizability across diverse datasets. In contrast, RFA-U-Net fully leverages RETFound’s large-scale self-supervised pretraining, enhancing its capability to detect intricate choroidal structures and boundary distinctions. By integrating a foundation model with an attention mechanism, RFA-U-Net achieves exceptional segmentation accuracy, particularly for boundary-sensitive tasks, while maintaining computational efficiency. While the RFA-U-Net has demonstrated superior performance, several limitations must be acknowledged. First, the model’s performance was validated on a single dataset, and its generalizability to other imaging modalities or populations remains to be explored. Additionally, the computational complexity introduced by attention gates and RETFound’s hierarchical encoding may present challenges in deployment on standard clinical hardware. Future research should focus on extending the model’s applicability to other retinal layers and integrating multi-task learning frameworks to simultaneously segment multiple layers or detect pathological features. Expanding the training dataset to include diverse populations and imaging devices will further enhance the robustness and clinical adoption of the model. Moreover, investigating lightweight or hardware-optimized versions of RFA-U-Net could facilitate its integration into point-of-care systems. The RFA-U-Net model’s robust segmentation capabilities can be extended to facilitate 3D reconstruction of the choroid layer. By segmenting consecutive OCT B-scans, the model can generate a series of accurately delineated 2D choroid layer masks, which can then be compiled into a volumetric representation. This approach enables detailed 3D visualization of the choroid, providing clinicians and researchers with critical insights into the structural and volumetric changes associated with various retinal diseases [44]. Such 3D reconstructions could further enhance diagnostic precision and enable dynamic tracking of disease progression or therapeutic responses over time. [45, 44]. In conclusion, this study introduced RFA-U-Net for choroid segmentation in OCT images, outperforming pre-trained CNN-U-Net architectures and SOTA methods. The integration of RETFound, a foundation model pre-trained on 1.6 million retinal images, enhanced feature extraction and generalization, while attention gates improved spatial selectivity. Additionally, Tversky loss addressed class imbalances, refining segmentation performance. These findings highlight the potential of foundation models and attention mechanisms in medical image segmentation, advancing precision and reliability in ophthalmology diagnostics.

## Data Availability

All data produced in the present study are available upon reasonable request to the authors corresponding authors

## V. DATA AVAILABILITY

Contact the corresponding authors

## VI. CODE AVAILABILITY

The code and models are publicly available at: https://github.com/Alirezahayatimedtech/RFA-U-Net.

